# Exploring Patients’ Needs & Desires for Quality Prenatal Care in Florida

**DOI:** 10.1101/2021.05.20.21257240

**Authors:** Kimberly Fryer, Chinyere N. Reid, Naciely Cabral, Jennifer Marshall, Usha Menon

## Abstract

**Introduction:** Early and adequate prenatal care is important for patients to obtain health education, risk reduction, psychosocial support, and necessary medical interventions during the prenatal period. High-quality prenatal care encourages adequate care throughout pregnancy by increasing patients’ desires to return for subsequent visits. This study aims to investigate the prenatal care experiences, desires, and needs of women in Florida.

**Methods:** A mixed-methods study was conducted from April to December 2019 with postpartum women (n=55) who received no or late prenatal care and delivered in [City], Florida. Eligible women completed a survey and a semi-structured interview. The interview contextualized conditions shaping perceptions of the quality of prenatal care. Univariate analysis was conducted on the demographic characteristics and prenatal care locations. Qualitative analysis was performed using MAXQDA software and modified grounded theory. The analysis was based on Donabedian’s quality of care model.

**Results:** The participants self-identified as Hispanic (52%), White (48%), and Black (21%). Sixty-nine percent had Medicaid while 22% were self-pay. In the qualitative analysis, three core themes emerged. Clinical care processes included provision of health education and medical assessments. Structural conditions included language barriers, clinic availability, and ancillary staff presence. Lastly, interpersonal communication included impersonal care and multiple providers. The overarching conclusion from this analysis was the desire for patient-centered care. Participants wanted the care they received to be informative, tailored to their needs, and to work within the realities of their everyday lives.

**Discussion:** Investigating women’s experiences with seeking and receiving prenatal care are necessary to inform future interventions. Fostering a positive and patient-centered healthcare environment is necessary to improve the quality of care.

**Quick points:** *What is already known on this subject?:*

- Racial and ethnic disparities exist in prenatal care access, utilization and both maternal and infant mortality and morbidity.
- Timely and adequate prenatal care is an important determinant for maternal and infant birth outcomes.

*What this study adds:*

- This study identified process, structure, and interpersonal factors associated with prenatal care access and utilization from the perspectives of a diverse group of postpartum women.
- Specific recommendations for patient-centered prenatal care for diverse women in the context of patient expectations and perceptions of quality are provided.

## Introduction

Maternal and infant morbidity and mortality are rising in the United States (US).^1^ Prenatal care offers an opportunity to screen, diagnose, and address risk factors and medical issues that may arise during pregnancy, with the goal of improving maternal and infant health outcomes. In the US, about 1 in 16 live births (6.2%) were to women who had late or no prenatal care, and about 1 in 7 live births (14.8%) were to women who had inadequate prenatal care.^2^ Late initiation of prenatal care and inadequate prenatal care have been found to be associated with preterm births, low birth weight, and neonatal mortality.^3–5^

While there has been a predominant focus on prenatal care adequacy, which consists of the timing of the first prenatal visit and the number of prenatal visits throughout pregnancy, limited research focuses on women’s perceptions of the quality of prenatal care and their preferences for prenatal care.^6–12^ The quality of prenatal care may be more crucial than prenatal care adequacy, especially to encourage women to participate. Therefore, inclusion of women’s perceptions of the quality of prenatal care they receive and preferences to meet their prenatal care needs, may potentially promote timely initiation and consistent prenatal care with the goal of improving infant and maternal health outcomes. This is particularly important in Florida which has a high number of undocumented women, uninsured women, and lower than average prenatal care initiation and adequacy rates ^13,14^. In this study, using Donabedian’s model of health care quality,^15^ we sought to assess how women in Florida perceive the quality of their prenatal care and to identify women’s preferences in prenatal care delivery to meet their needs.

## Methods

This study was conducted from April to December 2019 with 55 postpartum women who received late or no prenatal care and delivered at [hospital]. A retrospective mixed-methods study design explored participants’ views on the quality of their prenatal care. Participants completed open-ended surveys followed by semi-structured interviews that contextualized their survey responses to questions on prenatal care in terms of needs and desires. We used MAXQDA 2020 for analysis and codebook development.^16^ Donabedian’s conceptualization of health care quality^15^ as modified by Sword,^17^ informed the thematic analysis of the combined quantitative and qualitative data.

### Data Collection

Individuals eligible to participate were women over the age of 18 years who delivered a live-born infant, were able to speak or read either English or Spanish, and did not receive first-trimester prenatal care, defined as care prior to 14 weeks. We employed an identical sampling design whereby the quantitative and qualitative samples are the same.^18^ A purposive sampling strategy was used to recruit women on-site at the postpartum floor of one hospital from a list of participants whose eligibility was established through an initial EPIC electronic medical record (EMR) chart review.^19^

A total of 1,687 women were screened for eligibility through the initial chart review in EPIC EMR. Of the 210 individuals who both met the inclusion criteria and agreed to be approached by a member of the research team, 59 consented to participate in the study, and 4 were excluded due to not meeting inclusion criteria after enrollment. All participants signed informed consent to be enrolled in this study, which was approved by the University of [BLINDED] Institutional Review Board. We then implemented data collection as follows: 1) open-ended self-report survey administered to the participants by a member of the research team; 2) semi-structured interviews conducted to elaborate on participants’ meaning behind survey results, and 3) in-depth chart reviews of participants’ pregnancy characteristics. We collected the estimated date of delivery, delivery date, number of prenatal care appointments attended, and date of first prenatal care appointment from EMR chart reviews. Chart review data and survey responses were uploaded to Research Electronic Data Capture (RedCap).^20^ To understand participants’ perceptions of the quality of prenatal care received, participant characteristics and corresponding interview data were combined and coded using MAXQDA 2020 analytical software.^16^ Each participant received a $25 Wal-Mart gift card as compensation for participation.

#### Open-Ended Surveys

Open-ended surveys, administered in English or Spanish, included: 1) demographic questions about age, race, ethnicity, nativity, acculturation (Short Acculturation Scale for Hispanics),^21^ marital status, income, education level, parity, insurance type (before, during, and after pregnancy), and substance abuse during pregnancy; 2) pregnancy characteristics, clinic conditions and communication dynamics; and 3) an in-depth examination of the likes and dislikes related to prenatal care, and potential changes in prenatal care experience.

#### Interviews

Participants were asked a series of questions relating to their perception of their prenatal care experience (Table 1). Open-ended semi-structured interviews were audio-recorded by a member of the research team upon survey completion. Confidentiality concerns were addressed before initiating audio-recordings and participants were asked to refrain from using their names during the interview and told about the de-identification process.^19^ Almost all the participants consented to have their interviews audio-recorded (n=51). Three participants (i.e., ID 15, ID 55, and ID 58) declined audio-recording and detailed handwritten notes were taken during interviews.

**Table 1.**
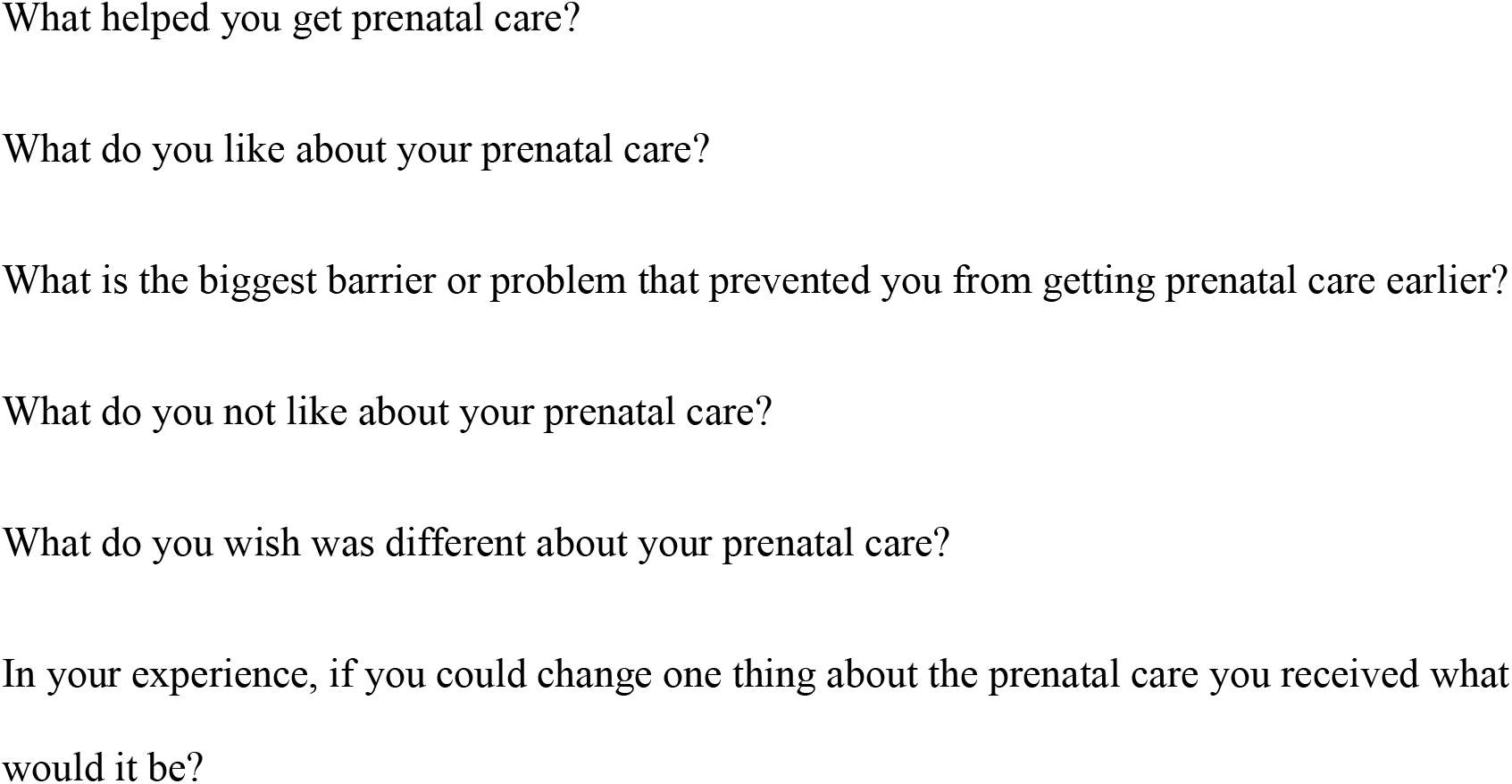
Open-Ended Survey Questions.

#### Quantitative Analysis

All chart review and survey data were entered into Redcap and analyzed with STATA (version 16). Sociodemographic variables were re-coded to facilitate univariate analysis of demographic characteristics. We conducted univariate analyses to examine sociodemographic characteristics of participants, and the prevalence of health, pregnancy, and health care measures in this population.

### Study variable manipulation

The following variables were re-coded to facilitate analysis: the substance abuse variable from the chart review; and, self-reported survey and interview data were collapsed into a variable that measures how many participants used any substances during pregnancy. The variables for nativity and ethnicity were collapsed and re-coded as yes/no to foreign-born (country of birth that is not the US or Puerto Rico). Years spent in school were re-coded and collapsed into categorical values: 1) some high school or less (0-11 years in school), 2) high school (12 years in school), 3) some college (13-15 years in school), and 4) college or higher education (≥ 16 years in school).

Spanish language interviews were translated directly from the Spanish recordings to English transcripts. These transcripts were then checked for accuracy by a native Spanish-speaking research assistant.

#### Qualitative Analysis

All interviews were audio-recorded and transcribed verbatim by a professional transcription service and potential identifiers were removed. During the first review, emergent codes were identified and then collapsed into subthemes. We used MAXQDA 2020 to support thematic coding. We drew on components of modified grounded theory (mGT) to identify key text segments using priori and in-vivo codes to develop a coding scheme ^22–24^. The code definitions were grounded in Donabedian’s evaluation of the quality of care^15^ as modified by Sword^17^ regarding how women view their prenatal care experience in terms of the structural conditions, clinical processes, and health outcomes they were subjected to while seeking or navigating clinic visits. We recorded analytic and structural memos to develop a codebook grounded in the theoretical framework.^24^ During later stages of this phase, earlier transcripts were reexamined for new codes, and the codebook was updated accordingly. Theoretical saturation was achieved in the analysis.

##### Validation strategy

Credibility for this study was achieved using the validation strategies of triangulation and peer debriefing.^19^ Data were triangulated between chart reviews, surveys, interviews, and field notes. Additionally, about a fifth (n = 10) of the transcripts were independently coded by another research team member to ensure reliability. Comparison of the inter-coder agreement computed for the first ten interviews was 92%. Coding discrepancies were identified and discussed to achieve consensus on the final codebook.

## Results

A total of fifty-five women took part in the study of mothers who had late or no prenatal care (n = 5; 9%) before delivering at [Hopsital]. Women were, on average, 29 years old (SD = 6.03) (Table 2). Most women preferred to communicate in English (n = 43; 78%); were born in the United States or Puerto Rico (n = 36; 67%); identified as White (n = 25; 45%), considered themselves Hispanic/Latinx (n = 28; 51%); and had a high school education or less (n = 38; 69%). Among those who affirmed “yes” to a Hispanic/Latinx heritage, most respondents self-identified as Mexican or of Mexican descent (43%). Moreover, twenty-two women were born outside the U.S. or Puerto Rico. Of the thirteen respondents who took the survey in Spanish, all used Spanish to speak, write, think, and interact with family and friends. On average, participants had their first prenatal care appointment 19 weeks (SD = 6.31) into their pregnancy; and went to eight prenatal care appointments (SD = 3.3). Although 22 women (41%) had no insurance a month before confirming their pregnancy status, 37 (67%) had Medicaid during delivery (Table 2). In this study, the most often used substance during pregnancy was marijuana (n = 12; 22%).

**Table 2.**
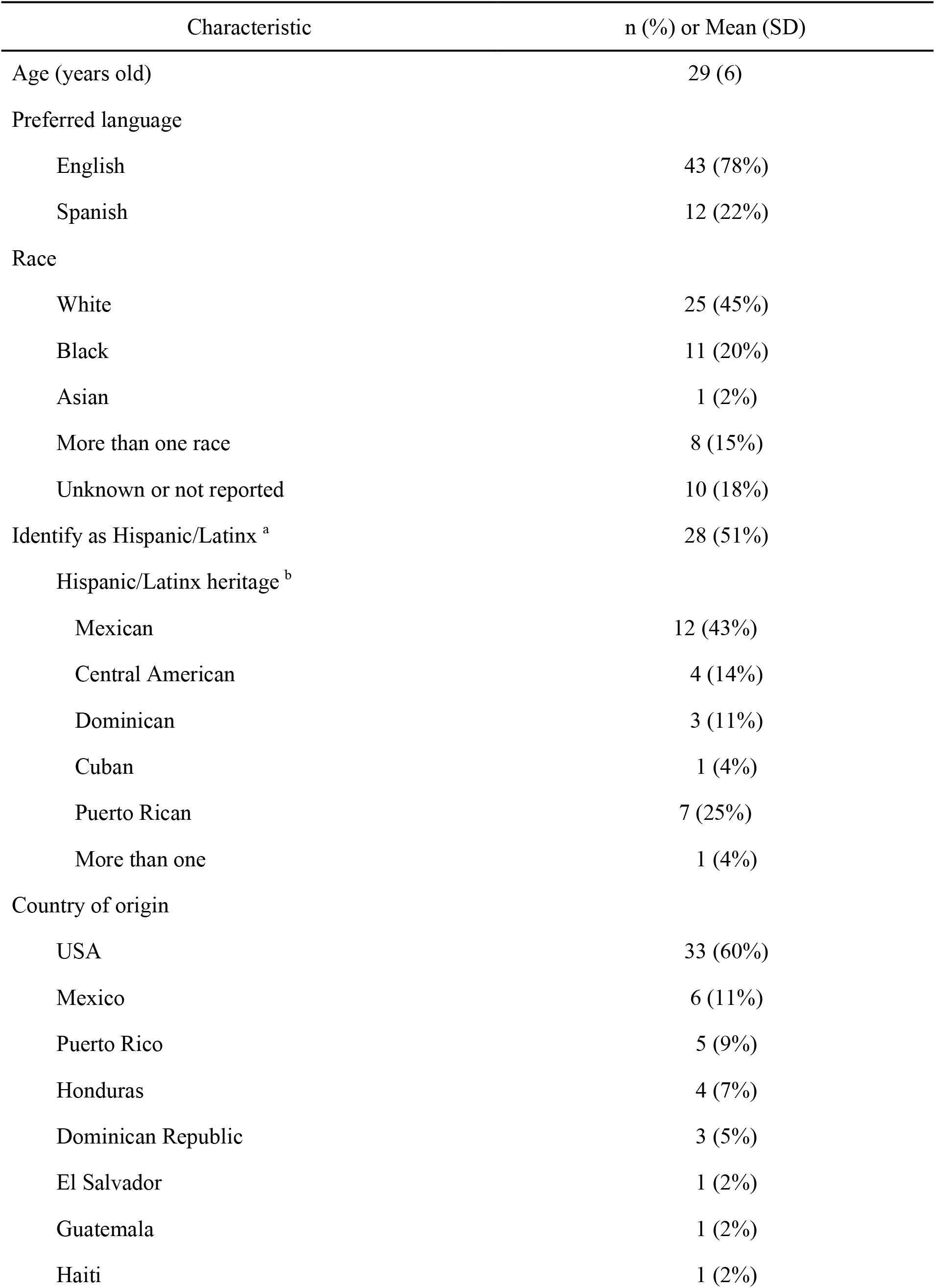
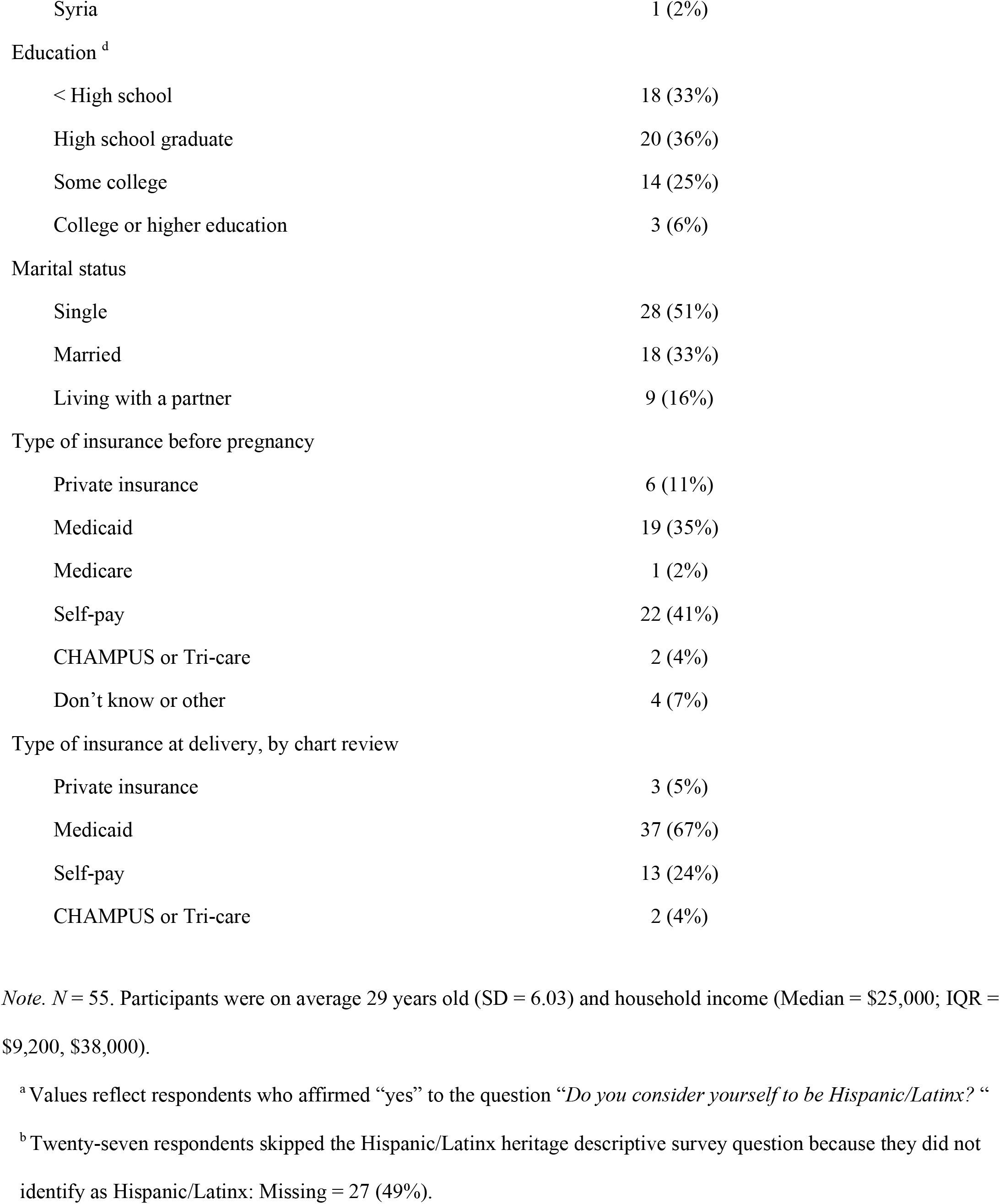
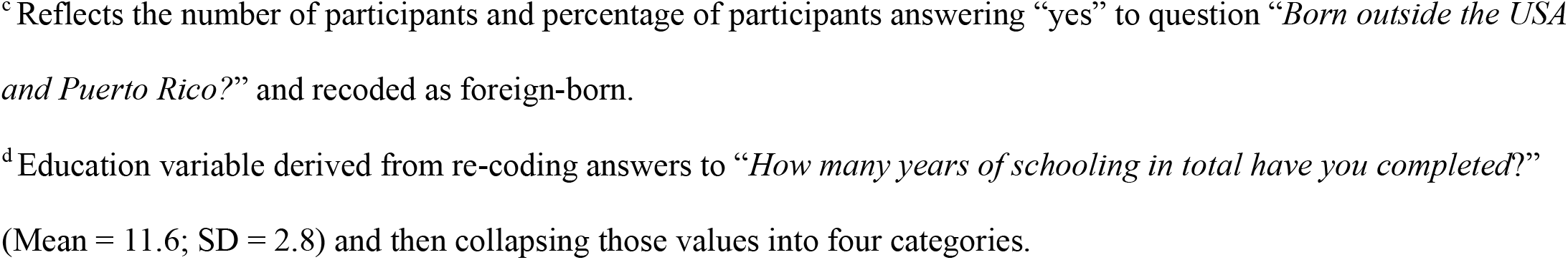
Sociodemographic Characteristics of women study participants (n = 55)

| Characteristic | n (%) or Mean (SD) |
| --- | --- |
| Age (years old) | 29 (6) |
| Preferred language |  |
| English | 43 (78%) |
| Spanish | 12 (22%) |
| Race |  |
| White | 25 (45%) |
| Black | 11 (20%) |
| Asian | 1 (2%) |
| More than one race | 8 (15%) |
| Unknown or not reported | 10 (18%) |
| Identify as Hispanic/Latinx <sup>a</sup> | 28 (51%) |
| Hispanic/Latinx heritage <sup>b</sup> |  |
| Mexican | 12 (43%) |
| Central American | 4 (14%) |
| Dominican | 3 (11%) |
| Cuban | 1 (4%) |
| Puerto Rican | 7 (25%) |
| More than one | 1 (4%) |
| Country of origin |  |
| USA | 33 (60%) |
| Mexico | 6 (11%) |
| Puerto Rico | 5 (9%) |
| Honduras | 4 (7%) |
| Dominican Republic | 3 (5%) |
| El Salvador | 1 (2%) |
| Guatemala | 1 (2%) |
| Haiti | 1 (2%) |
| Syria | 1 (2%) |
| Education <sup>d</sup> |  |
| < High school | 18 (33%) |
| High school graduate | 20 (36%) |
| Some college | 14 (25%) |
| College or higher education | 3 (6%) |
| Marital status |  |
| Single | 28 (51%) |
| Married | 18 (33%) |
| Living with a partner | 9 (16%) |
| Type of insurance before pregnancy |  |
| Private insurance | 6 (11%) |
| Medicaid | 19 (35%) |
| Medicare | 1 (2%) |
| Self-pay | 22 (41%) |
| CHAMPUS or Tri-care | 2 (4%) |
| Don't know or other | 4 (7%) |
| Type of insurance at delivery, by chart review |  |
| Private insurance | 3 (5%) |
| Medicaid | 37 (67%) |
| Self-pay | 13 (24%) |
| CHAMPUS or Tri-care | 2 (4%) |

*Note.* $N = 55$ . Participants were on average 29 years old ( $SD = 6.03$ ) and household income (Median = \$25,000; IQR = \$9,200, \$38,000).
<sup>a</sup> Values reflect respondents who affirmed “yes” to the question “*Do you consider yourself to be Hispanic/Latinx?*” “
<sup>b</sup> Twenty-seven respondents skipped the Hispanic/Latinx heritage descriptive survey question because they did not identify as Hispanic/Latinx: Missing = 27 (49%).
<sup>c</sup> Reflects the number of participants and percentage of participants answering “yes” to question “*Born outside the USA and Puerto Rico?*” and recoded as foreign-born.
<sup>d</sup> Education variable derived from re-coding answers to “*How many years of schooling in total have you completed?*” (Mean = 11.6; SD = 2.8) and then collapsing those values into four categories.

### Themes

Table 4 summarizes the main themes and sub-themes drawn from Donabedian’s model of quality of care^15^ as modified by Sword.^17^ Structure of care (at the clinic level), clinical care processes, and interpersonal communication were the three overarching themes that emerged from the data. In presenting the findings, we used supporting quotes to illustrate the patient’s perspectives in Table 4 represented by (theme acronym, quote #).

**Table 3.** Pregnancy Characteristics (N=55)

| Questions | Mean (SD) or percent<br>(%) |
| --- | --- |
| How many weeks pregnant were you when you were sure you were pregnant?, median (IQR) | 9 (4, 16)* |
| How did you feel when you found out you were pregnant with your new baby?* |  |
| I wanted to be pregnant later | 12 (22%) |
| I wanted to be pregnant sooner | 5 (9%) |
| I wanted to be pregnant at that time | 20 (37%) |
| I didn't want to be pregnant then or at any time in the future | 4 (7%) |
| I wasn't sure what I wanted | 13 (24%) |
| Way the pregnancy was dated |  |
| Ultrasound before 22 weeks | 26 (47%) |
| Ultrasound after 22 weeks | 28 (51%) |
| Last Menstrual Period only (no Ultrasound) | 1 (2%) |
| How long does it take you to get from your home to the clinic?, median (IQR) | 23 (13, 30)* |
| Did you get access to prenatal care as early in your pregnancy as you wanted?** |  |
| No | 35 (64%) |
| Yes | 20 (36%) |
| Gestational age at first prenatal visit by chart review in weeks, mean (SD) | 18.7 (6.3) |
| Number of prenatal care visits by chart review, mean (SD) | 7.5 (3.3)* |
| Gestational age at delivery by chart review, mean (SD) | 39 (2.43) |
| Any substance abuse during pregnancy? | 17 (31%) |

\* Missing = 1
\*\* Gestational age at delivery was calculated from Estimated Date of Delivery

**Table 4.** Quality of Prenatal Care Themes and Sub-Themes in Study with representative quotes.

| Theme and sub-themes | ID | Quotes |
| --- | --- | --- |
| **Structure of Care** |  |  |
| Clinic availability | SC1 | *“I wanted to go for a more natural vaginal birth, so I was looking for a provider that was more inclined to support that. I went with a doctor that offered midwifery or midwives... The options were very limited. I also considered a birthing center which I eventually switched over to and those are even more limited. Most of the midwives are associated with OB-GYNs and hospitals. So, there’s very few birthing centers, as well.” (ID 38-Hisp)* |
|  | SC2 | *“What I don’t like? I would say it wasn’t in a timely fashion for me to set an appointment...I didn’t think that it would take so long if I would’ve scheduled it at Genesis that I would be seen like two months later...I just didn’t know who to contact.” (ID 25-NHB)* |
|  | SC3 | *“So, we had scheduled our appointment. I had found out when I was eight weeks and it [appointment] wasn’t scheduled until 13 weeks, and when we went to our first appointment, I hadn’t been accepted by Medicaid yet. So, it would’ve been a lot of money out of pocket. So, we ended up having to reschedule it and then waiting until we got accepted for Medicaid.” (ID 28-Hisp)* |
|  | SC4 | *“Well... I went back and looked, and my first ultrasound was actually February 4th, so that was like eight weeks. But that was just like a confirmation of pregnancy before they would start me in prenatal care, and then that was a month later, so that’s why I ended up like being 12 or 13 [weeks].” (ID 36-NHW)* |
| **Clinical Care Processes** |  |  |
| Screening and medical assessments | CCP1 | *“[Knowing] the status of the baby if he’s growing right, if he has any defects going on. Me, to see if I have any health issues while I’m carrying the child because your body can* |
|  |  | *change. It's just basically knowing about my health while I'm pregnant. That's about it."*<br>(ID 25-NHB) |
|  | CCP2 | *"Like I wanted to know ... like what will happen with the baby, like if they can see in the ultrasound. But they didn't have that, they just had like checking the heartbeat and that's it. They just sent me here-- like the next appointment...they just did the ultrasound, that was the only time I did have an ultrasound. But I wish I could've seen more of my baby even before anything." (ID 12-Hisp)* |
|  | CCP3 | *"Absolutely. I was left on a medication called Lamictal...a folic acid antagonist, so what that means is it increases the chances of my child having spina bifida, and because of this, I was denied at two OB-GYN clinics...I wish that both of the original clinics that denied me would have been able to inform me on the first phone call that they would not accept a patient who had these risk factors and would have been able to give me a clear direction to someone who would accept somebody in my situation. Also, I would have wished that they would have understood how simple it would be to deem me not high risk because all it took was an ultrasound for them to deem me fine, that my baby was safe. I wasn't in harm's way. There was nothing that would have deemed this pregnancy high-risk. It would have been a very simple task. That, I wish is what I could have changed." (ID 29-NHW)* |
|  | CCP4 | *"Uh \*exhale\*, the prescription process, whenever I went and they would change my – my prescriptions or, you know, they start you out on the prenatal vitamins all the time, like every time I got a prescription it wasn't able to be filled like at my pharmacy that I go to...I think it would have been more convenient if I'm going to this clinic one or two times a week that they have a pharmacy on site that carry the medications that are prescribed for pregnant females." (ID 18-Hisp)* |
| Health education concerns | CCP5 | *"...so, they had done a blood test. It was—I thought that it was for the baby. They looked into it and found out I had CML, which is chronic myeloid leukemia and other than that? They have been great helping me learn about the baby and how to be a mom. It's my first* |
|  |  | *time with a kid... They actually helped me a lot thorough the process of it—becoming a mom and going through my depression stage because of the medicines that I take...*<br>*Depression was like a huge thing in my pregnancy that I was scared that she [the baby] was going to catch it. So, they uh, come in and sat me down and we talked about every two to three times a month about it. So, it made me feel a lot better, knowing that they cared like that. It's that I don't really like taking pills, so the fact they actually sat me down and talked about it was really helpful. I mean, I have family at home, but they are working, and they don't really have time for that—they got this and this to do. So, with them, they just sat me down and we talked about it. And they helped me understand more about the pregnancy.” (ID 54-More than one race)* |
|  | CCP6 | *“So, I was a first-time mom so I didn't know a bunch of the risks and all that and stuff like that - like that needed to be... I knew that you couldn't take like certain pain killers and stuff like that but not in depth.” (ID 28-Hisp)* |
|  | CCP7 | *“To me, it seemed like every time you go there, they tell you the same information over and over again. So, I feel like they need to do other stuff within your visits... To me, it's just they tell you the weight and the same stuff that you know every time. I think that they need to do more within the visits.” (ID 05-NHB)* |
|  | CCP8 | *“Through Medicaid. I had done research. I went ahead and looked online myself and seen what all our Medicaid covers and what they provide so that I could benefit from every little thing that they have because I knew I was going to need everything that I can get. There weren't any pamphlets that notified me of these things. These are things that I had to do on my own, and that's another barrier. I feel like there should be more outreach program and more people get out giving knowledge about - making people knowledgeable about what health benefits they have access to because the lack of knowledge is like the biggest problem. People aren't knowledgeable about the care that they can receive and so they're not getting it” (ID 47-NHB)* |

| Interpersonal Communication |  |  |
| --- | --- | --- |
| Impersonal care | IC1 | *‘Well, what I would change about the clinic, of course the doctors...the doctor that was there at first was rude. I don’t like it when you’re rude and disrespectful and just act like you don’t care. This is your job. This is what you have to do. You know what you basically signed up for and you know what you’re going to come here to do. If you don’t want to do your job, why are you here?’ (ID 33-Hispanic)* |
|  | IC2 | *“The staff from the other two clinics treated me very poorly based on the judgment that I was a bad mother for getting pregnant while on a medication that could cause something as detrimental to my child as spina bifida. They treated me very unkindly and that was probably the biggest factor that I did not like. They judged me very heavily and it was a feeling I would never like to repeat for myself or anyone else...Then at the second office, the office staff, they are very judgmental. Thankfully, the doctor was not judgmental, but they were very unkind. They did not wish to speak to me any further about things such as, “Why would you not have gotten care before this?” The second clinic stated that, “How could you go so long without care,” which I informed them that I had previously been refused...” (ID 29-NHW)* |
|  | IC3 | *“I just thought they were always in a rush because they have other people to assess...I wish like they would’ve told me more about what was going on.” (ID 12-Hisp)* |
|  | IC4 | *“We see too many different doctors. I don’t like that because I feel like they don’t know me personally and they don’t know what’s going on or how I feel...You can tell which doctor is very interested in you and your care. The doctors are like 10 minutes and then it’s like, “Okay, see you later,” out the door...You can see the same doctor or at least three doctors instead of 10 doctors. Also, the doctors that you see, should be one of the doctors that deliver your baby because I see all these different doctors, but that wasn’t even the doctor that delivered my baby. I don’t even know her. I’ve never even seen her... [so] you are* |
|  |  | *nervous because you don't know anything about her. You really can't trust a person you don't know." (ID 40-Hisp)* |
| Positive relationship dynamics | IC5 | *"It was just like the staff was really - they made me feel comfortable. Also, when I came in, they knew exactly who I was and they made sure I was checked in properly, made sure I was in my appointment on time. They made sure that I didn't stay over any longer, and they were just all nice, smiling, and every time I left, they told me, "Goodbye. Have a nice day. See you later." No one was rude or mean or anything... They didn't make me feel uncomfortable and make me not want to go to my appointments. I made it to mostly every appointment..." (ID 24-NHB)* |
|  | IC6 | *"They made me feel actually safe and they broke everything down to make sure I understood like what was going on and just like the plans that they have for me and during the pregnancy." (ID 20-Hisp)* |
|  | IC7a | *"They treated me well--excellently—very well when they took care of me. One experience that I liked? The nurses were excellent with me. They showed me how I was doing in my pregnancy...When I left, I felt fine. They treated me very well." (ID 58-Hisp)* |
|  | IC7b | *"I really like the midwives because they're much more understanding than the doctors. They usually take a lot more time to be with each patient and address their concerns. They're usually much more open to actually discussing issues or procedures...one of the midwives, which I actually saw her for most of my appointments there...She took the time to make sure that my medications were working for my mental health and just – there was one appointment where I just sat there and complained to her about all the things going wrong in my life [Laughter] and she just listened..." (ID 51-Hisp)* |
|  | IC8 | *"I liked the prenatal care and the visits...I've been going to them since I had my first kid, which was I was like 17, so they're like my only experience really. All the doctors are really nice. Everybody is really sweet, you know what I mean? Like they take really good care of you; I love that about it. When the doctors do see you, they make sure they go over* |
|  |  | *like everything, like if I have any concerns, they make sure they address every single concern. They don't really like push me off or rush me out, you know what I mean?" (ID 23-NHB)* |
| Language barriers | IC9 | *"Communicating with the nurses is hard when you don't speak English." (ID 19-Hisp)* |
|  | IC10 | *"When I came here for the first time for the ultrasound, I did not understand much of what they were telling me about the ultrasound. I told the doctor, but he was rude, very blunt, when he said to me that the baby might come sick. I mean, what I understood from him was that it [the baby] was coming sick.... I do not know if it was him that did not explain it well or if I understood it wrong)." (ID 11-Hisp)* |

\**Hisp* – Hispanic, *NHB* – non-Hispanic Black, *NHW* – non-Hispanic White

### Structure of Care

Structural conditions frame environments of care, which in turn influence how individuals perceive access to and quality of the care available to them. The relevant sub-theme included attitudes to clinic availability.

#### Clinic availability

The sub-theme of clinic availability encompasses attitudes and lingering desires among women who could not schedule their first prenatal care visit earlier in their pregnancy or subsequent prenatal visits. Most participants complained of long wait times to be scheduled for their first and subsequent prenatal visits because clinics were fully booked, and this was attributed to several factors. Particularly, Black, and Hispanic women reported limited and inconvenient clinic hours that were not compatible with their schedule, and the unavailability of preferred providers or clinics when they wanted to schedule their appointments (SC1). However, based on the perceived quality of care and rapport with doctors, most of these participants opted to wait until their desired clinic became available. Additionally, Black and Hispanic women experienced difficulties navigating who to talk to about and how to schedule an appointment (SC2). The time taken to receive Medicaid and the acceptance of Medicaid by providers was also associated with delays in scheduling the first prenatal appointment especially for Hispanic participants and an Asian participant (SC3). Furthermore, a White participant attributed the requirement of the clinic that her pregnancy be confirmed before scheduling her first prenatal visit to be the cause of her delay in prenatal care (SC4). Overall, participants reported long wait times experienced during prenatal visits as a negative experience of prenatal care.

### Clinical Care Processes

Clinical care processes refer to the application of medical knowledge and clinical procedures implied in the provision of care.^15^ Relevant sub-themes within clinical care are 1) screening and medical assessments, and 2) health education concerns. Interviews prompted discussions on attitudes and lingering desires for health promotion, information sharing, clinical testing, and medical assessments expected during prenatal care.

#### Screening and medical assessments

This sub-theme frames attitudes towards screening tests and health assessments, and lingering concerns about the types of knowledge and health assessments available to pregnant women resulting in later prenatal care. Overall, participants across all races desired to know the health status of their baby and themselves during pregnancy through screenings and medical assessments. Participants reported that awareness of their baby’s health status was mainly by hearing the baby’s heartbeat and having several ultrasounds done throughout the pregnancy, attributing this to quality of prenatal care (CCP1). However, some women reported only hearing their baby’s heartbeat but not receiving what they regarded as sufficient ultrasounds to ensure the health of their baby (CCP2). Perceived barriers to receiving these ultrasounds were described by White participants as being a result of lacking health insurance and being scheduled for tests and ultrasounds separately from regular prenatal appointments. Similarly, blood draws though understood to be necessary, were of concern to White participants, who complained that they were uncomfortable, and results were not discussed in a timely manner. The ability to address maternal health issues and fears were expressed by participants across all races. For example, an Asian participant discussed how her elevated blood pressure was mismanaged, while another White participant expressed how her fears related to past drug use and mental health issues were not addressed appropriately (CCP3). On the other hand, the availability of prescribed medication at the clinic where prenatal care was being received was of importance to minority participants, (CCP4).

#### Health education concerns

Transparency and information on treatments and services during prenatal care appointments improve perceptions of approachability to health care services during pregnancy.^25^ Generally, all participants valued and desired health information. Health information concerns were related to pregnancy, prenatal care, Medicaid, and access to health services. Some participants reported receiving desired information and having concerns related to pregnancy addressed. For example, a multiracial participant who was diagnosed with Chronic Myeloid Leukemia during prenatal screening found her providers helpful in educating her about her diagnosis, medication, and depression associated with her illness (CCP5). However, despite some participants’ positive experiences with receiving information on their health and available services, participants across racial groups felt uninformed about the clinical care processes. For instance, participants desired more information about disease risk factors, medication risks in pregnancy, nutritional information, services offered, and desired for information to be delivered in a clear manner (CCP6). Participants also preferred to have access to information earlier and did not like receiving repetitive information at each visit (CCP7). Although some minority women relied on Medicaid as a source of information for health and services, some had difficulty comprehending Medicaid benefits and coverage (CCP8).

### Interpersonal Communications

Interpersonal communication dynamics reflect negative and positive interactions with providers that encourage or discourage participants in seeking or staying in prenatal care.

#### Impersonal care

Impersonal care encompassed the negative interactions and relationships encountered with healthcare providers when receiving prenatal care. Provider professionalism and lack of continuity of care with the same provider were related to quality of prenatal care. Lack of provider professionalism described as the provider being rude (IC1), dismissive, or being judgmental was reported by participants across all races, except by Black participants. For example, a White participant felt judged by staff at several clinics regarding her pregnancy situation, thereby affecting how she viewed the quality of prenatal care she received (IC2). Also related to provider professionalism was the desire by minority participants to not feel rushed or juggled around during their prenatal visits (IC3).

An emphasis on the lack of continuity of prenatal care providers was perceived negatively towards quality of prenatal care. Participants across all races valued seeing the same provider at each prenatal visit and attributed one-on-one doctor-patient visits as more personal, relating this to building trust and rapport with their provider (IC4).

#### Positive relationship dynamics

Positive relationships include efforts to promote health and knowledge sharing in a positive and welcoming environment. Generally, participants reported that providers and staff being nice, friendly, and supportive fostered a positive environment for prenatal care. For a Black participant this positive environment encouraged her adherence to prenatal care visits (IC5). Likewise, providers who were considered understanding and shared needed information in a comprehensible manner were valued by participants receiving prenatal care (IC6). However, Hispanic women perceived themselves as having a more positive relationship dynamic with nurses compared to doctors because nurses were deemed more helpful and understanding (IC7a). Similarly, this sentiment was also shared by a White participant (IC7b). As described by a Black participant, a history of building a relationship with a provider from prior pregnancy prenatal visits reinforced continued positive relationship dynamics with the same provider (IC8).

#### Language barriers

Language barriers (in this case Spanish) hinder one’s ability to understand the information shared. The language barriers sub-theme encompassed language-related issues in obtaining prenatal care through communication with providers and clinical staff. Some Spanish-speaking participants had difficulty communicating with clinic staff in English (IC9), while for some it was not an issue because clinic staff spoke Spanish. Equally important, language barriers might also contribute to confusion and miscommunication of health information for women seeking prenatal care (IC10).

#### Synthesis of Findings

After synthesizing the responses, the overarching conclusion from this analysis was the desire for patient-centered care. Patient-centered care is care which is consistent with the values and preferences of patients and includes shared decision making between the provider and patient (Figure 1).^26,27^ Participants wanted the care they received to be informative, tailored to their needs, and work within the realities of their everyday lives. When care was instead focused on the needs of the healthcare system instead of the patients, the quality of care suffered. For example, rigid clinic schedules and long wait times due to limitations imposed by the healthcare system did not meet the needs of the patients and was not patient-centered care. When a participant found a provider who took time to explain their care plan and focus on their emotional needs, the care was considered high quality. Not having access to providers who could speak Spanish or interpreters interfered with patients’ understanding of care and communication. Interpersonal dynamics were an important feature called out by patients, which may be even more critical for women of color.

**Figure 1:**
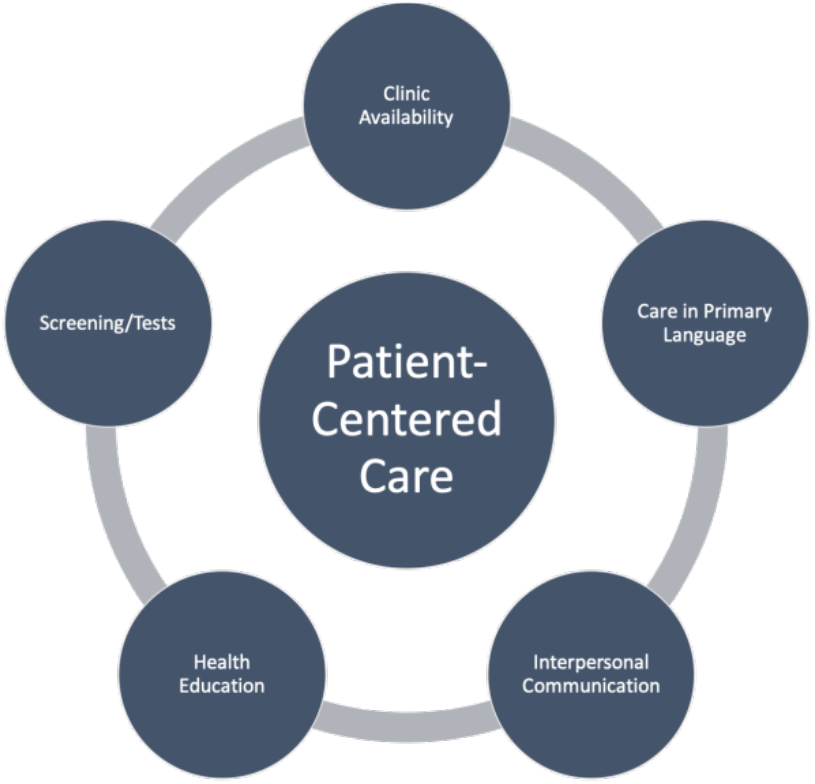
Patient centered care.

## Discussion

Using Donabedian’s model of health care quality,^15^ we sought to assess how women in Florida perceive the quality of their prenatal care and to identify women’s preferences in prenatal care. We found that women wanted care which included screening and medical assessments, provided health education, included easy clinic access, and was offered their native language with providers who were personal and supportive. These elements align with the framework of patient-centered care.^26,27^

Compared to research by Sword in Canada,^17^ our participants wanted similar elements in prenatal care. Both populations desired providers that interacted with them with a respectful attitude, took the time to talk with them and did not rush through the visit, and provided emotional support. Both populations also valued sharing of health information, screenings and assessments, and continuity of care. Unlike in the research by Sword et.al, our participants noted more issues with scheduling such as long wait times for appointments and for being seen once in clinic, which may be related to the different healthcare delivery systems in each study. We also noted several issues with access; those themes and analysis are presented more fully in a separate paper.^28^ Due to our diverse sample, we also found that language barriers for Spanish-speakers were an important element to effective prenatal care.

This research provides new elements for further improvement in prenatal care and modifiable factors in the clinical setting. We noted many areas for quality improvement in the provision of prenatal care which could be incorporated into new models of prenatal care or improve existing models. For example, the predominant model of care, individual prenatal care, could be improved through promoting interpersonal care, relationship building and modifying scheduling to allow for provider continuity. Additionally, adding after-hour clinical appointments or more flexible appointment scheduling could improve care and increase uptake of prenatal care visits. Women of color experienced particular issues with competing life demands that precluded fitting with clinic times, but health education concerns were experienced more evenly across groups.

These elements of quality prenatal care also create patient-oriented outcomes for new models of prenatal care such as virtual prenatal care, group prenatal care or enhanced prenatal care models. Overall, while some Black and Hispanic participants may have reported different issues than White, our findings are not generalizable. As such, further research is needed to delve deeper into each of these elements to determine measurable outcomes, but ensuring that future evaluations of models of care include patient-centered outcomes will allow for improved quality of care and prenatal care that women want to access and utilize during their pregnancy. It is also imperative that we better understand if specific needs of women of color are different from those of white women beyond the overt such as language. For example, cultural nuances of beliefs related to pregnancy and pregnancy outcomes may need to be incorporated once understood in more detail, however, these were not identified in our study. These patient-oriented outcomes can be combined with the research by Peahl,^10^ on patient preferences for prenatal care delivery to develop enhanced models of prenatal care that are patient-centered.

Our study included several strengths and weaknesses. Participants were very diverse and included non-English speakers. As many models of care and studies on prenatal care have excluded non-English speakers,^10,29,30^ we sought to include these women and other low-income women of color as these subgroups have the highest maternal mortality in the United States.^31^ Limitations include recall bias and a single-site design. As we approached women postpartum, we relied on their recall of their prenatal care experiences. Our sample was from one, large tertiary care hospital. We acknowledge that women in different communities may have different preferences for prenatal care, and all models and methods of prenatal care must be tailored to each community.

## Conclusions

In our study, perceptions and expectations of quality prenatal care were aligned with the constructs of patient-centered organizational structures, clinical care, and interpersonal communication. The assessment of barriers to unique specific minority groups should be extended to better understand cultural nuances and avoid stereotyping women seeking prenatal care. To improve early and sufficient access to prenatal care for diverse patients, providers and organizations should attend to the ease in which patients access care appointments, align the services provided during the visit in the context of patient expectations, and practice effective interpersonal communication and relationship-building.

## Data Availability

Data Use Agreement with the University of South Florida is required for data access.

